# THE EFFECT OF A BOARDING RESTRICTION PROTOCOL ON EMERGENCY DEPARTMENT CROWDING

**DOI:** 10.1101/2021.05.01.21255822

**Authors:** Ji Hwan Lee, Ji Hoon Kim, Incheol Park, Hyun Sim Lee, Joon Min Park, Sung Phil Chung, Hyeon Chang Kim, Min Joung Kim

## Abstract

**Background:** Access block due to a lack of hospital beds causes emergency department (ED) crowding. We initiated the “boarding restriction protocol” that limits ED length of stay (LOS) for patients awaiting hospitalization to 24 hours from arrival. This study aimed to determine the effect of the protocol on ED crowding.

**Method:** This was a pre-post comparative study to compare ED crowding before and after protocol implementation. The primary outcome was the red stage fraction with more than 71 occupying patients in the ED (severe crowding level). LOS in the ED, treatment time and boarding time were compared. Additionally, the pattern of boarding patients staying in the ED according to the day of the week was confirmed.

**Results:** Analysis of the number of occupying patients in the ED, measured at 10-minute intervals, indicated a decrease from 65.0 (51.0–79.0) to 55.0 (43.0–65.0) in the pre- and post-periods, respectively (p<0.0001). The red stage fraction decreased from 38.9% to 15.1% of the pre- and post-periods, respectively (p<0.0001). The proportion beyond the goal of this protocol of 24 hours decreased from 7.6% to 4.0% (p<0.0001). The ED LOS of all patients was similar: 238.2 (134.0–465.2) and 238.3 (136.9–451.2) minutes in the pre- and post-periods, respectively. In admitted patients, ED LOS decreased from 770.7 (421.4–1587.1) to 630.2 (398.0–1156.8) minutes (p<0.0001); treatment time increased from 319.6 (198.5–482.8) to 344.7 (213.4–519.5) minutes (p<0.0001); and boarding time decreased from 298.9 (109.5–1149.0) to 204.1 (98.7–545.7) minutes (p<0.0001). In the pre-period, boarding patients accumulated in the ED on weekdays, with the accumulation resolved on Fridays; this pattern was alleviated in the post-period.

**Conclusions:** The protocol effectively resolved excessive ED crowding by alleviating the accumulation of boarding patients in the ED on weekdays. Additional studies should be conducted on changes this protocol brings to patient flow hospital-wide.

## INTRODUCTION

Worldwide, emergency department (ED) crowding is a serious public health problem.[1, 2] It is well known that ED crowding causes various adverse effects such as increased mortality, increased medical errors, ambulance diversion, and financial loss suffered by medical institutions.[3, 4] “Access block” refers to the inability to transfer a patient from the ED to a hospital bed, thus the patient is boarded in the ED, even after the emergency treatment has been completed.[5, 6] Access block, one of the main contributors to ED crowding, occurs when hospital bed occupancy increases.[5, 7] Alan et al. reported that the length of stay (LOS) of admitted patients in the ED increased by 18 minutes for every 10% increase in hospital bed occupancy, and increased dramatically when bed occupancy exceeded 90%.[8]

To solve this problem, several countries have implemented mandatory national policies limiting the time spent by patients in the ED. In 2004, the UK’s National Health Service first introduced the “4-hour target” that required 98% of patients who visited the ED to leave within 4 hours of arrival.[9] Since then, similar national policies have been initiated in several countries. The Australian government applied the National Emergency Access Target (NEAT), which limited the ED LOS to 4 hours as in the UK, and New Zealand adopted a longer 6-hour target.[10, 11] The governments of these countries had a strong will to improve ED crowding and demanded this improvement from hospitals rather than from EDs alone, and these mandatory policies had a great effect on improving the indicators of ED crowding.[12-13] In Korea, ED crowding has been a long-standing and serious public issue, but there is no national regulation on ED LOS. The Korean government did not actively intervene in the improvement of crowding and just recommended a permissible threshold (below 5%) for keeping patients in the ED for more than 24 hours. It is difficult to solve the access block problem because it requires changing hospital rules regarding bed arrangements, but hospital leadership will not be easily motivated to change the rules in the absence of government policy.

We initiated a new protocol, the “boarding restriction protocol,” to control the situation in which patients who needed to be hospitalized wait in the ED for unlimited periods of time due to hospital crowding. The core content was to limit the LOS of each patient waiting for hospitalization to 24 hours from their arrival at the ED. We aimed to study the effect of this boarding restriction protocol on ED crowding.

## METHOD

### Study design and setting

This was a pre-post comparative study conducted in the ED of a tertiary university hospital located in an urban area. The hospital had approximately 2,200 beds, with an average annual bed utilization rate of 80% or more. The ED was divided into an adult ED and a pediatric ED; the adult ED, where this study was conducted, consisted of a monitoring area (13 beds), a bed area (28 beds), a chair area (20 recliners), and a fast track area. The adult ED treated patients over 16 years, and the number of visiting patients was about 90,000 per year. About 20% of visiting patients were hospitalized, and the average waiting time for hospitalization was 10 hours. Patients who stayed in the ED for more than 24 hours was 5–10% of total patients. Emergency medicine doctors commenced treatment for all patients arriving in the ED, and if hospitalization was deemed necessary, consultation with the relevant specialist was conducted to determine whether the patient should be hospitalized. When hospitalization was indicated, the attending physician took over the patient’s treatment, and treatment continued in the ED until a hospital bed was ready. When a patient was required to be transferred to another hospital, this was arranged by a request to the ED coordinator. The boarding restriction protocol began in November 2019. However, coronavirus disease-19 (COVID-19) infections began to spread in Korea from February 2020, which greatly affected the ED’s treatment process. Therefore, the duration of this study was 9 weeks, starting from the first week of December 2019, and was compared with the same period one year previously. All data were collected by the hospital information system and processed anonymously, and ethical approval was waived from Yonsei University Health System, Severance Hospital, Institutional Review Board (no. 4-2020-1164).

### Boarding restriction protocol

The boarding restriction protocol is intended for patients requiring hospitalization and the purpose of the protocol is to remove these patients from the ED within 24 hours of their arrival (Figure 1). The strategy to achieve this goal was to allocate hospital beds within 18 hours from ED arrival. If the bed assignment was not made within 18 hours, a text message was sent to the attending physician informing them that the patient could not be admitted due to the absence of a hospital bed. When the attending physician decided to transfer the patient to another hospital, the ED coordinator arranged an appropriate hospital according to the patient’s condition and dispatched the patient within 24 hours. If the patient could not be transferred to another hospital for reasons such as their severe condition or refusal to be transferred, the attending physician could decide to keep the patient in the hospital. In the event that the patient was kept in the hospital, that patient was given bed allocation priority over the other patients waiting in the attending physician’s outpatient clinic.

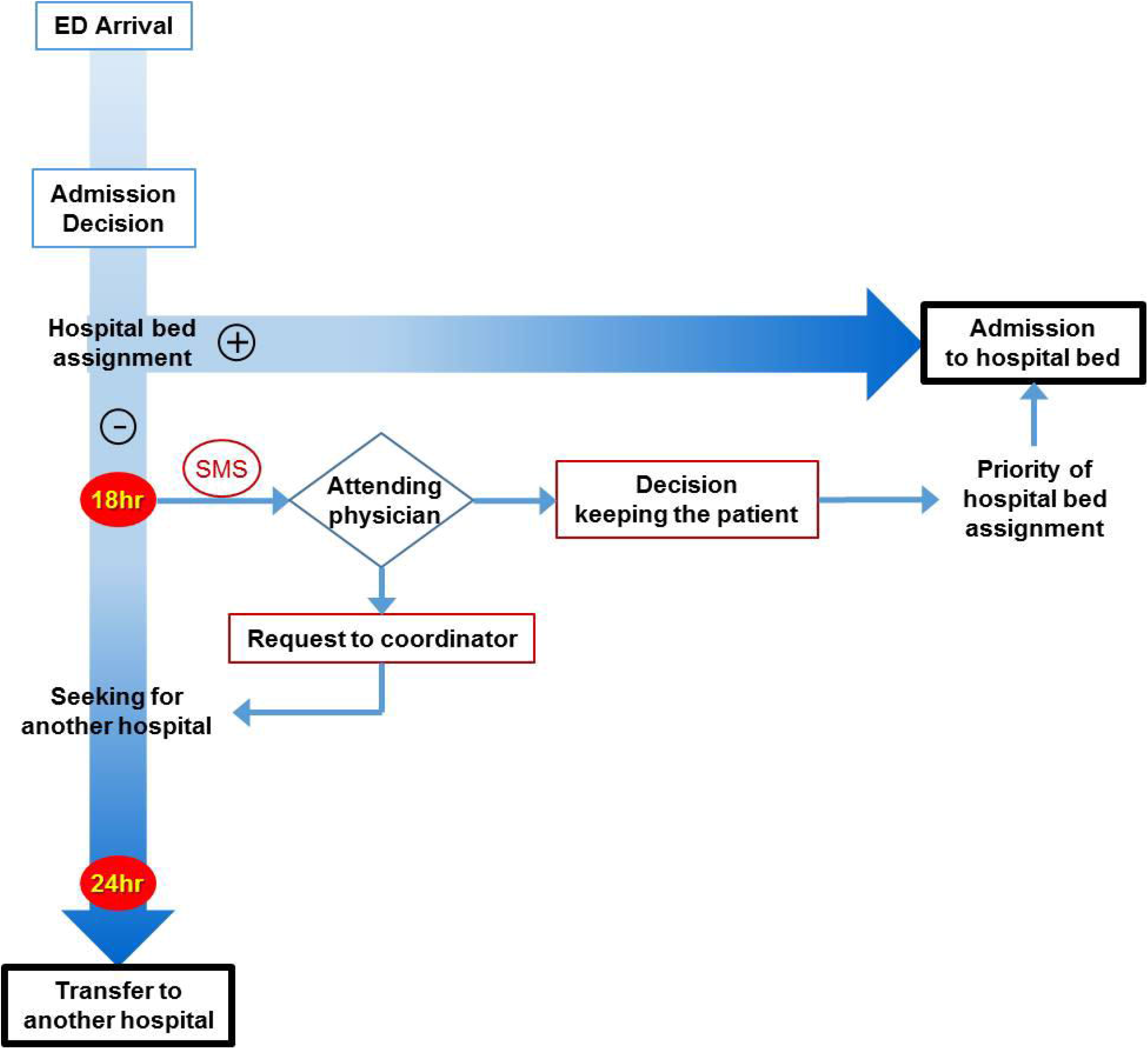

### Crowding indicator

In the ED, the crowding status was monitored and divided into three stages based on the number of occupying patients: green for 50 or less, yellow for 51 to 70, and red for 71 or more. The green stage meant that the ED was operating smoothly without crowding, and the yellow stage meant that patient treatment could not proceed smoothly due to moderate crowding. The red stage meant that it was difficult to treat new patients due to severe crowding. As the primary outcome of this study, the change in the red fraction, before and after protocol implementation, was confirmed. To determine the fraction of the crowding stage, we reconstructed the dataset of the number of ED patients at 10-minute intervals from the time of arrival to the time of departure of each patient. The secondary outcome was the proportion of patients who stayed longer than 24 hours and the ED LOS. Time factors were also investigated according to the ED treatment process. The “treatment time” was the time taken from ED arrival to the disposition decision (admission, discharge, or transfer); the decision time for transferred patients was based on the time when the doctors made a transfer request to the coordinator; and the “boarding time” was defined as the time from the decision of disposition to the time of leaving the ED.

Since the target of this protocol was boarding patients, their LOS and occupancy in the ED were analyzed in detail. The ED occupancy by patients decided to be hospitalized, but still staying in the ED, was reconstructed at 10-minute intervals using each patient’s admission decision time and ED departure time. Since hospitalization patterns are very different on weekdays and weekends, the number of patients admitted to hospital beds and ED LOS were analyzed according to the day of the week.

We also checked the occupancy rate of hospital beds and the total number of hospitalized patients in both periods. Hospital bed occupancy was calculated at eight o’clock every morning as the number of occupying patients out of the total number of beds.

### Study variables

Patient data were extracted from the hospital information system and electronic medical records. We checked whether the patient was transferred from another hospital and whether they arrived via emergency medical services. The result of triage by Korean Triage and Acuity Scale (KTAS), a five-point classification scale (1=resuscitation, 5=non-emergent), was investigated. The complaint category was the organ system that corresponded to the main symptom presented by the patient. Non-medical problems referred to ED visits due to external issues such as trauma, poisoning, or environmental factors. Severe disease was determined based on the severe disease group designated by the Central Emergency Medical Center under the Ministry of Health and Welfare. We investigated whether the emergency physician treated the patient and the area where the treatment commenced. Time of ED arrival and weekend were classified based on the time the patient arrived at the ED and whether it was a weekend or not. Laboratory study, imaging study (x-ray, computed tomography, magnetic resonance imaging), and specialty consultation performed during the ED stay were also confirmed.

### Statistical analysis

Nominal variables were compared between the pre- and the post-period using the chi-square test and presented as a number with a percentage. Variables of time factors were analyzed by Mann-Whitney U test considering the positive skewness of the data distribution, and were shown as median and interquartile range (IQR). The crowding stage was classified from the number of ED occupying patients at 10-minute intervals, and the proportion of each stage during the study period was compared between the two groups. A logistic regression for patients staying longer than 24 hours and a generalized linear regression for ED LOS was performed by selecting influencing variables with p-values less than 0.05 in univariable analysis. The analysis was performed using SAS (version 9.4, SAS Inc, Cary, NC, USA) and was determined to be statistically significant when the p-value was less than 0.05.

## RESULTS

### Patient characteristics

A total of 12,498 patients during the pre-period and 13,050 patients during the post-period were treated in the ED (Table 1). The sex and age of both groups were similar, and the proportion of patients who were transferred from other hospitals decreased from 12.9% in the pre-to 10.8% in the post-period (p<0.0001). The proportion of less urgent patients with KTAS 4 and 5 decreased during the post-period, and the proportion of patients diagnosed with severe disease increased from 17.8% to 18.9% (p=0.0313). The rates of laboratory study and specialty consultation were similar between the two periods, but among the imaging study, CT was performed more in the post-period (37.5% vs. 38.9%, p=0.0202). As a result of treatment, the number of hospitalized patients decreased from 2,853 (22.8%) to 2,793 (21.4%), and the number of patients transferred to other hospitals increased from 347 (2.8%) to 399 (3.1%) (p=0.0128).

**Table 1.** Comparison of patient characteristics between pre- and post-period

| Variable |  | Pre-period<br>n=12,498 | Post-period<br>n=13,050 | p-value |
| --- | --- | --- | --- | --- |
| Age, n (%) | –39 | 3897 (31.9) | 4151 (31.8) | 0.9761 |
|  | 40–64 | 4502 (36.0) | 4697 (36.0) |  |
|  | 65– | 4009 (32.1) | 4202 (32.2) |  |
| Female, n (%) |  | 6719 (53.8) | 6930 (53.1) | 0.2925 |
| Transfer in, n (%) |  | 1612 (12.9) | 1404 (10.8) | <0.0001 |
| EMS, n (%) |  | 2995 (24.0) | 3144 (24.1) | 0.8106 |
| KTAS, n (%) | 1 | 200 (1.6) | 201 (1.5) | <0.0001 |
|  | 2 | 960 (7.7) | 1172 (9.0) |  |
|  | 3 | 2651 (21.2) | 3798 (29.1) |  |
|  | 4 | 6857 (54.9) | 6508 (49.9) |  |
|  | 5 | 1830 (14.6) | 1371 (10.5) |  |
| Complaint category, n (%) | Gastrointestinal | 2498 (20.0) | 2538 (19.5) | 0.8055 |
|  | General | 2086 (16.7) | 2242 (17.2) |  |
|  | Neurological | 1815 (14.5) | 1844 (14.1) |  |
|  | Cardiovascular | 1290 (10.3) | 1388 (10.6) |  |
|  | Musculoskeletal | 1084 (8.7) | 1119 (8.6) |  |
|  | Respiratory | 993 (8.0) | 1014 (7.8) |  |
|  | ENT | 818 (6.6) | 845 (6.5) |  |
|  | Skin | 794 (6.4) | 843 (6.5) |  |
|  | Other | 1120 (9.0) | 1217 (9.3) |  |
| Severe disease, n (%) |  | 2234 (17.8) | 2469 (18.9) | 0.0313 |
| Emergency physician, n (%) |  | 5433 (43.5) | 5238 (40.1) | <0.0001 |
| Area, n (%) | Monitoring area | 1000 (8.0) | 922 (7.1) | <0.0001 |
|  | Bed area | 2101 (16.8) | 2497 (19.1) |  |
|  | Chair area | 2093 (16.8) | 2754 (21.0) |  |
|  | Fast track | 7304 (58.6) | 6877 (52.7) |  |
| Time of ED arrival, n (%) | 0–6 | 2265 (18.1) | 2329 (17.9) | 0.3466 |
|  | 6–12 | 3531 (28.3) | 3807 (29.2) |  |
|  | 12–18 | 3786 (30.3) | 3858 (29.6) |  |
|  | 18–24 | 2916 (23.3) | 3056 (23.4) |  |
| Weekend, n (%) |  | 4131 (33.1) | 4061 (31.1) | 0.0009 |
| Laboratory study, n (%) |  | 8892 (71.2) | 9237 (70.8) | 0.5197 |
| Imaging study, n (%) | X-ray | 9428 (75.4) | 9955 (76.3) | 0.1135 |
|  | CT | 4680 (37.5) | 5071 (38.9) | 0.0202 |
|  | MRI | 679 (5.4) | 688 (5.3) | 0.5680 |
| Specialty consultation, n (%) |  | 6960 (55.7) | 7275 (55.8) | 0.9254 |
| Treatment result, n (%) | Admission | 2853 (22.8) | 2793 (21.4) | 0.0128 |
|  | Discharge | 9298 (74.4) | 9858 (75.5) |  |
|  | Transfer | 347 (2.8) | 399 (3.1) |  |

SD: standard deviation; EMS: emergency medical services; KTAS: Korean Triage and Acuity Scale; ED: emergency department; CT: computed tomography; MRI: magnetic resonance imaging.

### ED crowding

Analysis of the number of occupying patients in the ED, measured at 10-minute intervals, indicated a decrease by 15.4% from 65.0 (51.0-79.0) in the pre-period to 55.0 (43.0-65.0) in the post-period (p<0.0001). Figure 2 shows the distribution of the number of occupying patients according to the day and time of the week. Overall, the number of ED patients in the post-period decreased throughout the week compared to in the pre-period. The pattern of crowding resolving itself at dawn and worsening in the afternoon was observed in both periods. Crowding gradually worsened from Monday to Thursday in the pre-period, but this was not observed in the post-period. The red stage fraction, indicating severe crowding, decreased from 38.9% in the pre-period to 15.1% in the post-period (p<0.0001).

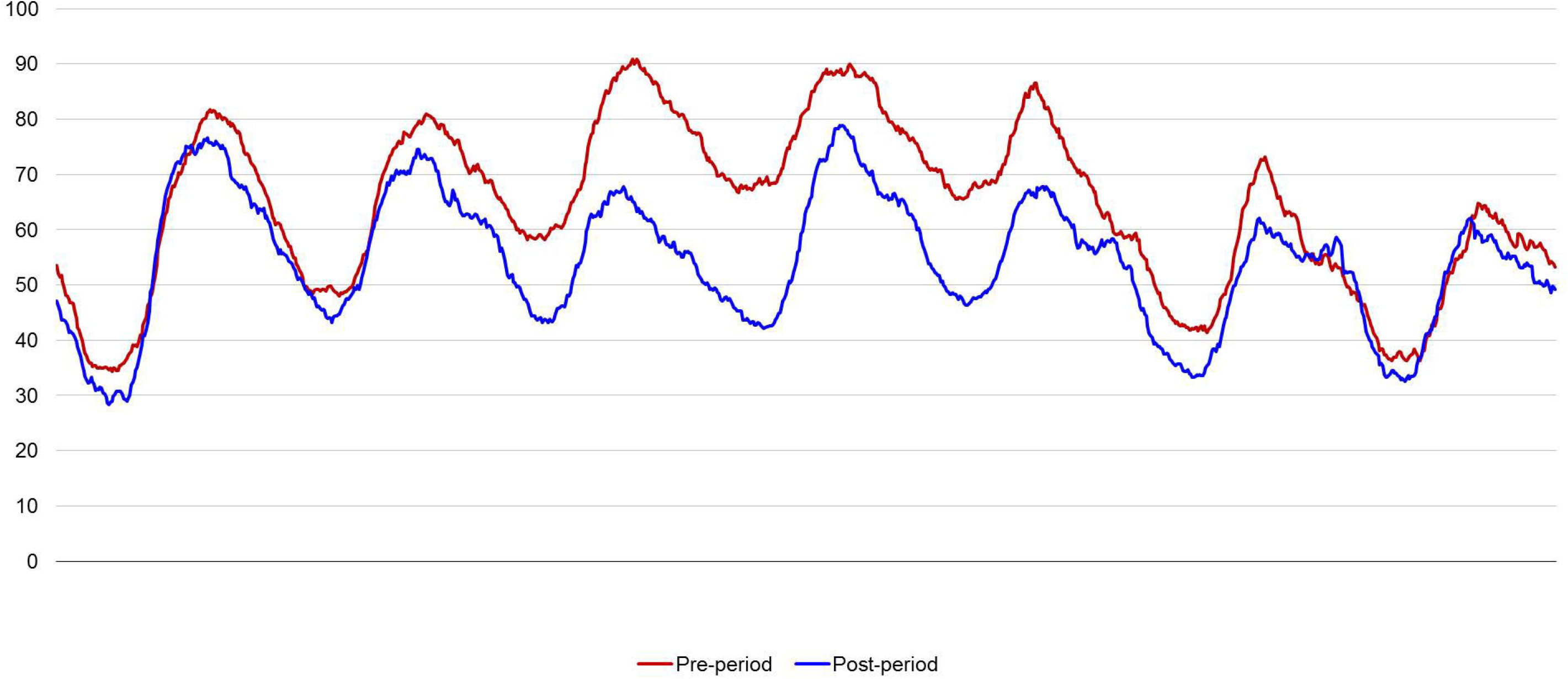

The number of patients leaving the ED beyond the protocol’s goal of 24 hours decreased from 951 (7.6%) to 525 (4.0%) (p<0.0001). The results of logistic regression analysis for the proportion of patients with LOS exceeding 24 hours are shown in Appendix 1. After correcting influencing factors, the effect of the protocol on LOS over 24 hours was statistically significant with an odds ratio (OR) of 0.433 (0.384–0.489). In admitted patients, the post-period had an adjusted OR of 0.428 (0.372–0.492) for patients staying for over 24 hours (Appendix 2).

The ED LOS of all patients was 238.2 (134.0–465.2) minutes in the pre-period and 238.3 (136.9–451.2) minutes in the post-period (Table 2). After adjusting for influencing factors, the post-period was a significant factor for ED LOS with a beta coefficient of −99.3 (−113.1 to −85.4) (Appendix 3). In admitted patients, ED LOS decreased from 770.7 (421.4–1587.1) minutes to 630.2 (398.0–1156.8) minutes (p<0.0001); with a beta coefficient of −310.9 (−360.6 to −261.3) (Appendix 4). Treatment time increased by 7.9% from 319.6 (198.5–482.8) minutes to 344.7 (213.4–519.5) minutes (p<0.0001), while boarding time decreased by 31.8% from 298.9 (109.5–1149.0) minutes to 204.1 (98.7–545.7) minutes (p<0.0001). The ED LOS of transferred patients increased from 379.1 (255.8–695.5) minutes to 443.8 (278.3–695.5) minutes, and the boarding time increased by 50.2% from 93.3 (55.1–164.6) minutes to 140.1 (89.4–226.9) minutes.

**Table 2.**
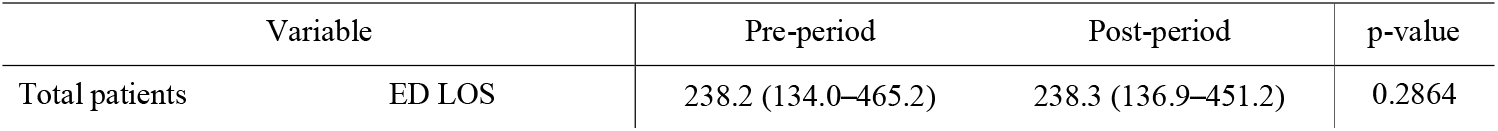
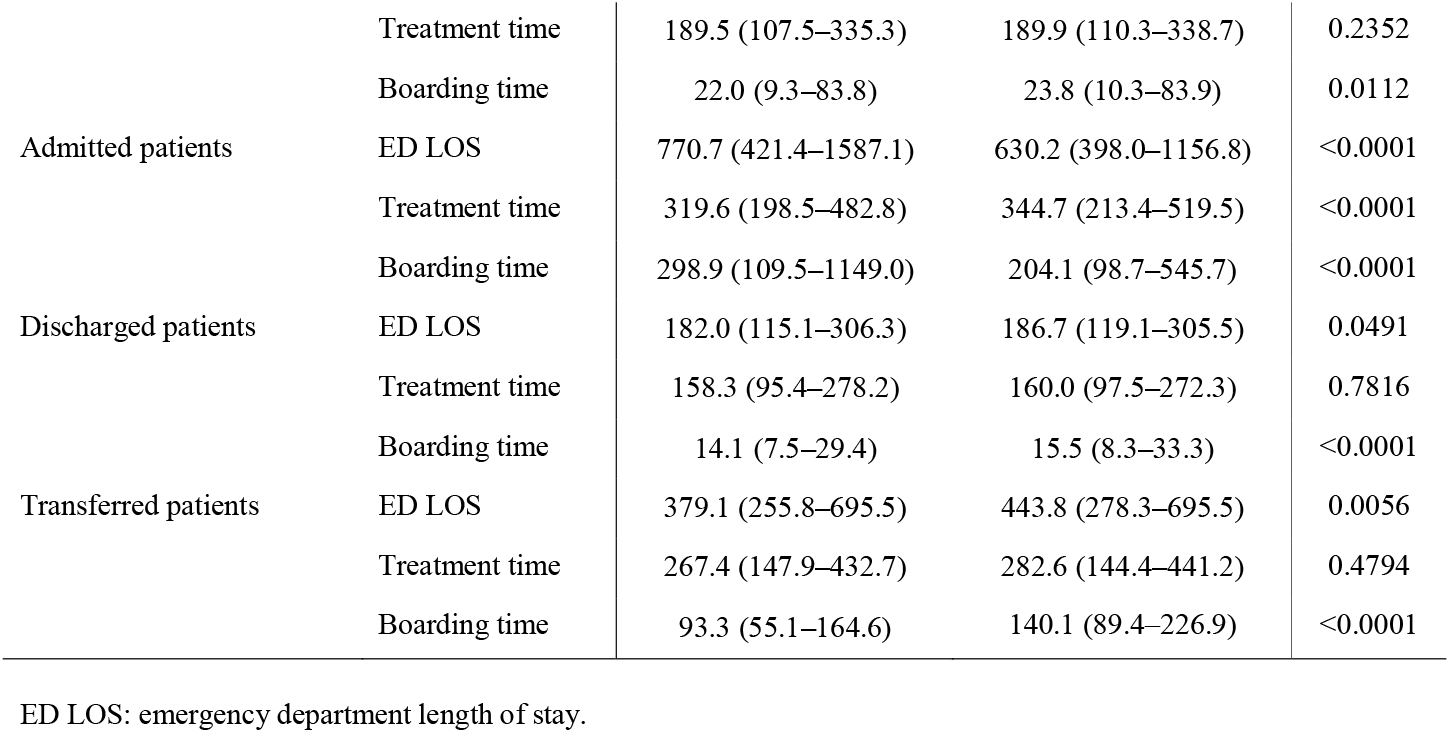
Comparison of emergency department length of stay and time factors between pre- and post-period

### Boarding patients in the ED

Figure 3 shows the distribution of the arrival and departure days of the admitted patients by day of the week. During the pre-period, the number of patients admitted to hospital beds was smaller than the number of patients who arrived at the ED from Monday to Thursday, maintaining a positive difference, and from Friday onwards, more patients could be hospitalized. During the post-period, the number of patients arriving and leaving on Tuesday and Wednesday remained similar, and hospitalization was less concentrated on Friday compared to in the pre-period. In the pre-period, the ED LOS of patients arriving from Tuesday to Thursday was significantly delayed, but this feature was alleviated in the post-period.

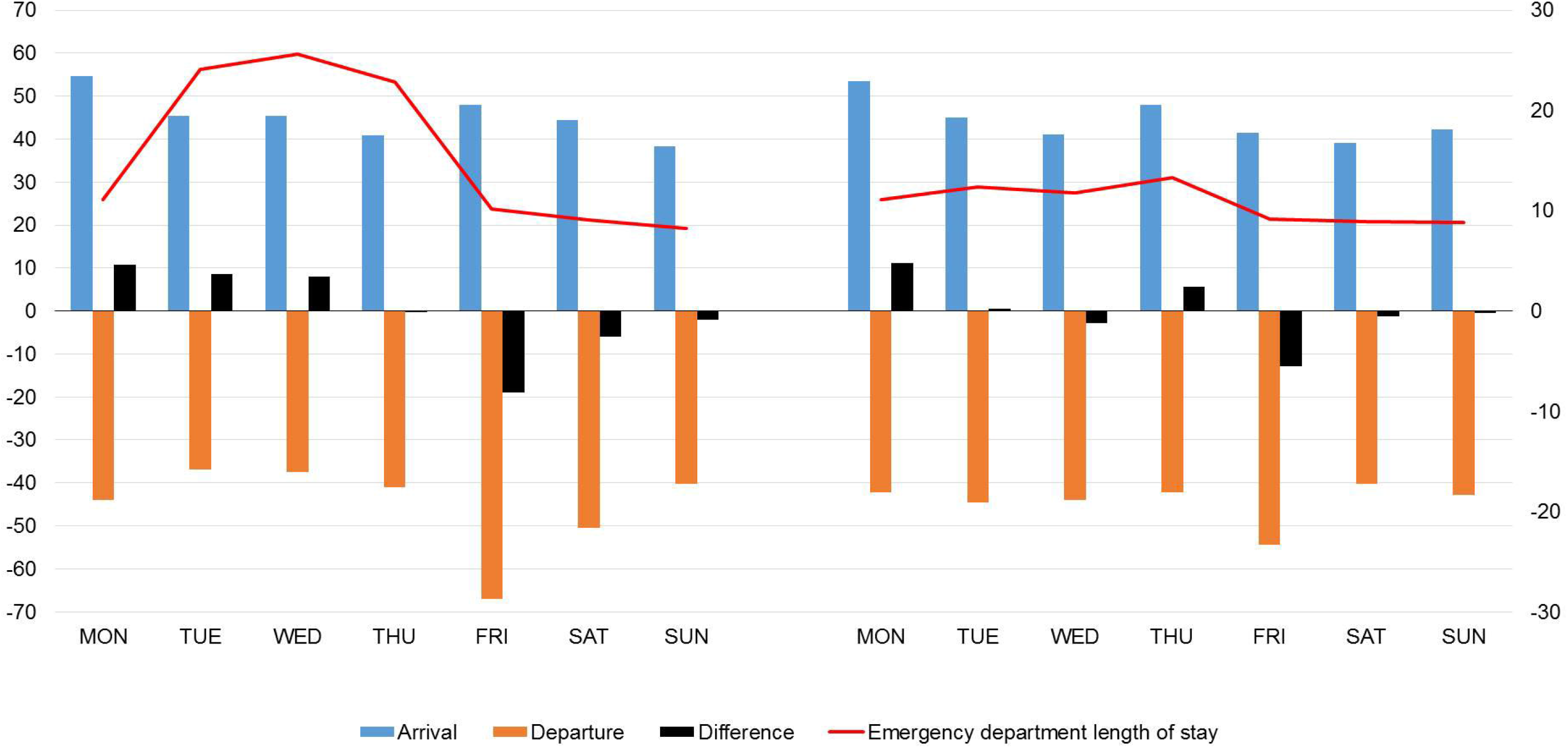

The median number of boarding patients measured at 10-minute intervals was 21.0 (12.0–37.0) in the pre-period and 10.0 (7.0–19.0) in the post-period, thus showing a decrease of 52.4% (p<0.0001). In the distribution of boarding patients, the number of patients continuously increased from Monday to Thursday and then rapidly decreased on Friday in the pre-period (Figure 4). The number of boarding patients during the post-period was also higher on weekdays than on weekends, but the difference was less than in the pre-period.

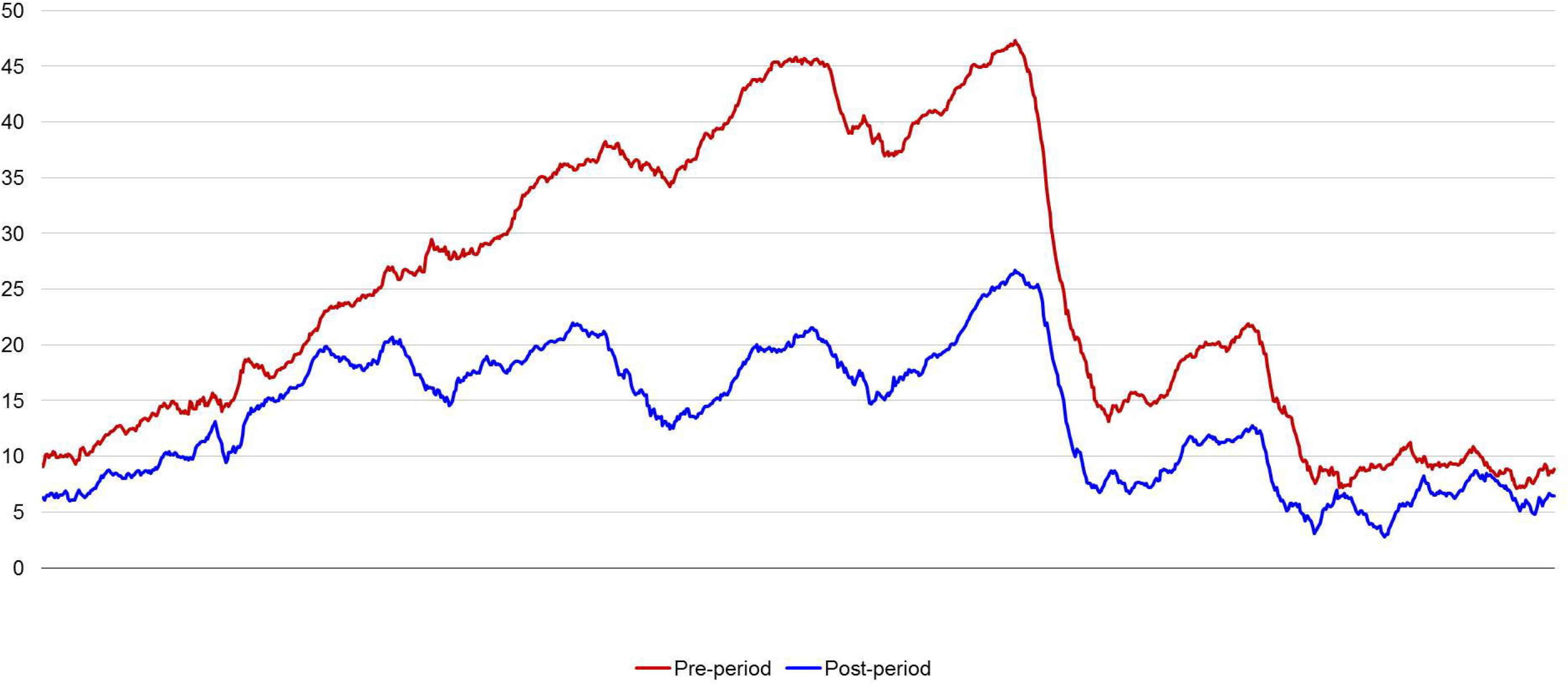

The total number of patients admitted to the hospital was 21,626 in the pre-period and 21,258 in the post-period. Among them, 2,839 (13.1%) and 2,794 (13.1%) patients were admitted via ED, respectively, and there was no statistically significant difference between the two periods (p=0.9619). The hospital bed occupancy in both groups was similar with 86.3 (81.2–87.6) in the pre-period and 83.8 (79.2–88.8) in the post-period (p=0.3609).

## DISCUSSION

In this study, we confirmed that the boarding restriction protocol significantly reduced the red stage fraction in the ED by reducing the LOS of admitted patients. Although only 21.4% of the patients were hospitalized, it was surprising that managing their LOS within 24 hours could reduce the extreme overcrowding situation from 38.9% to 15.1%. It is well known that the main driver of ED crowding is obstruction of outflow.[14, 15] Since access block is caused by crowding of the entire hospital, the solution should not be limited to the ED, and bed capacity must be increased in consideration of patient flow throughout the hospital.[15, 16] However, there is still not much incentive for leadership to recognize ED crowding as a hospital-wide problem, and this can also be related to the hospital’s profits; thus, it is not easy to change the hospitalization policy of the entire hospital to solve ED crowding. For this reason, we could not proceed with the entire hospital and had to start with only emergency patients inside the ED, even though our protocol aimed to solve the output of the ED. Even so, our protocol would have affected the patient flow throughout the hospital, because we not only pushed emergency patients out of the hospital, but also gave the attending physician the authority to decide to admit emergency patients prior to those in their outpatient clinics.

As discharged patients accounted for 75.5% of total patients and were not subject to this protocol, the entire ED LOS was not reduced. In admitted patients, we were able to observe an interesting phenomenon: while ED LOS decreased as boarding time was significantly reduced, treatment time increased. It is understood that admission decision-making took longer because the attending physician had to be more cautions when making decisions as patients could not board indefinitely in the ED until hospitalization. This finding is in line with previous studies that increased ED crowding was associated with a decrease in decision-making regarding hospitalization.[17] ED crowding is a desperate situation for emergency staff as it hinders the provision of adequate first aid to new emergency patients, but the attending physicians outside the ED are not directly affected by ED crowding. Until this protocol was initiated, the attending physicians made admission decisions without considering the availability of hospital beds, and bed allocation was the responsibility of the administrative staff. Since administrative staff could not consider the medical condition of patients, patient safety was inevitably threatened when hospital beds were insufficient. With this protocol, attending physicians were forced to face the problem of hospital bed shortages and intervene in the assignment of beds, so that emergency patients could have bed priority.

We observed that there was congestion in the number of patients waiting for hospitalization in the ED on weekdays and that this congestion was resolved on Friday and Saturday; this pattern was resolved considerably after the implementation of the protocol. A significantly longer ED LOS of admitted patients who arrived at the ED on Tuesday, Wednesday, and Thursday was also resolved. Since scheduled hospitalization is mainly conducted on weekdays, the delay in emergency hospitalization during weekdays has been reported in previous studies.[7, 18, 19] In most hospitals, medical staff prefer to perform procedures at the beginning of the week and to discharge patients before the weekend. This preference is because doctors do not want to work on the weekends, but if the patient is still in the hospital over the weekend, they have to continue to care for the patients.[20] The variation in the hospital’s daily inpatient census is a combination of the natural variation of emergency hospitalization and the artificial peak and valley of scheduled hospitalizations. The discrepancy between the hospital’s available resources and patient demand is a major cause of the degradation of quality of care, which impedes access to care, and ultimately threatens the safety of patients.[21, 22] To efficiently use hospital resources and ensure patient safety, the peaks and valleys of patient demand must be smoothed.[23-25] The demand smoothing interventions introduced in previous studies were accompanied by system changes such as weekend staffing relocation to alleviate the weekend effect.[16. 23, 24, 26] Although this protocol did not directly intervene in artificial variation, it achieved smoothing of the weekly variation of hospitalization through the ED by limiting boarding time in the ED. Further studies should be conducted on the effect of smoothing of the weekly fluctuation in emergency admission on patient flow throughout the hospital and on patient safety. Since it is difficult to cancel scheduled surgeries and procedures, hospitalization of patients who need conservative treatment may be delayed. Excessive delay in scheduled hospitalizations may also be a factor that hinders patient safety from a long-term perspective, such as delayed chemotherapy. Thus, a system that proactively coordinates overall hospitalizations is essential.

The present study has some limitations. First, because our study has a pre-post comparative design and was performed retrospectively, some confounders may not have been identified. Second, to avoid the COVID-19 outbreak period that had a major impact on ED processes, a short study period of 9 weeks was unavoidable. Thus, it was impossible to ascertain the long-term effects of this protocol. Finally, this study was conducted in a single tertiary hospital with a high bed occupancy of more than 80% and a significantly long boarding time in the ED. Therefore, the effect of this protocol may be different in hospitals where the degree of crowding and the pattern of patient flow is different. It is expected that the protocol will be effective in hospitals where boarding in the ED is severe as a result of access block.

## CONCLUSION

In this study, it was confirmed that the boarding restriction protocol was effective in resolving excessive ED crowding. This was possible because weekly variations in emergency hospitalization were alleviated by facilitating the hospitalization of emergency patients on weekdays. Further research is needed on the changes made to patient flow throughout the hospital and the impact of these changes on patient safety and hospital revenue. Furthermore, it is necessary to establish a system to control and optimally operate the input and output of all patients admitted throughout the hospital, whether for emergency or elective treatments.

## Data Availability

Data are available upon reasonable request.

## Notes

### Competing Interest Statement

The authors have declared no competing interest.

### Funding Statement

This work was supported by the National Research Foundation of Korea grant funded by the Korea government (Ministry of Science and ICT) (no. 2017R1C1B5076958).

### Author Declarations

All data were collected by the hospital information system and processed anonymously, and ethical approval was waived from Yonsei University Health System, Severance Hospital, Institutional Review Board (no. 4-2020-1164).

