## Appendix for "THE EFFECT OF A BOARDING RESTRICTION PROTOCOL ON EMERGENCY DEPARTMENT CROWDING"

Appendix 1. Logistic regression analysis for emergency department length of stay over 24 hours in total patients.

| Variables | | Univariable | | Multivariable | |
| --- | --- | --- | --- | --- | --- |
|  |  | Odds ratio (95% CI) | p-value | Odds ratio (95% CI) | p-value |
| Age | -39 | 1 | <0.0001 | 1 |  |
|  | 40-64 | 3.737 (3.108-4.493) |  | 1.638 (1.344-1.995) | <0.0001 |
|  | 65- | 5.574 (4.653-6.676) |  | 1.448 (1.186-1.768) | 0.0003 |
| Female |  | 0.579 (0.520-0.644) | <.0001 | 0.728 (0.648-0.818) | <0.0001 |
| Transfer in |  | 2.398 (2.112-2.724) | <.0001 | 1.018 (0.882-1.174) | 0.8113 |
| EMS |  | 2.314 (2.078-2.578) | <.0001 | 1.120 (0.975-1.286) | 0.1086 |
| KTAS | 1 | 1.660 (1.249-2.207) | 0.0005 | 1.535 (1.074-2.195) | 0.0187 |
|  | 2 | 1.103 (0.939-1.294) | 0.2327 | 0.982 (0.809-1.192) | 0.8549 |
|  | 3 | 1 |  | 1 |  |
|  | 4 | 0.359 (0.318-0.405) | <.0001 | 0.720 (0.612-0.849) | <0.0001 |
|  | 5 | 0.198 (0.153-0.255) | <.0001 | 0.688 (0.510-0.927) | 0.0142 |
| Complaint category | Gastrointestinal | 1 |  | 1 |  |
|  | General | 0.725 (0.596-0.882) | 0.6966 | 1.404 (1.178-1.673) | 0.0002 |
|  | Neurological | 0.098 (0.056-0.170) | <.0001 | 0.233 (0.182-0.298) | <0.0001 |
|  | Cardiovascular | 0.970 (0.830-1.133) | 0.0013 | 0.552 (0.443-0.687) | <0.0001 |
|  | Musculoskeletal | 0.300 (0.223-0.402) | <.0001 | 0.556 (0.402-0.768) | 0.0004 |
|  | Respiratory | 0.526 (0.433-0.639) | <.0001 | 1.570 (1.313-1.876) | <0.0001 |
|  | ENT | 0.349 (0.266-0.457) | <.0001 | 0.402 (0.225-0.717) | 0.0020 |
|  | Skin | 2.618 (2.240-3.060) | <.0001 | 0.928 (0.545-1.582) | 0.7845 |
|  | Others | 0.130 (0.080-0.212) | <.0001 | 0.476 (0.355-0.637) | <0.0001 |
| Severe disease |  | 4.374 (3.928-4.870) | <.0001 | 1.545 (1.362-1.754) | <0.0001 |
| Emergency physician |  | 3.678 (3.278-4.127) | <.0001 | 1.150 (0.987-1.341) | 0.0740 |
| Area | Monitoring area | 4.887 (4.207-5.676) | <.0001 | 1.396 (1.123-1.736) | 0.0027 |
|  | Bed area | 3.080 (2.711-3.499) | <.0001 | 1.276 (1.085-1.500) | 0.0032 |
|  | Chair area | 0.838 (0.698-1.006) | 0.058 | 0.674 (0.553-0.821) | <0.0001 |
|  | Fast track | 1 |  | 1 |  |
| Time of ED arrival | 0-6 | 1 |  | 1 |  |
|  | 6-12 | 3.369 (2.724-4.167) | <.0001 | 2.285 (1.796-2.906) | <0.0001 |
|  | 12-18 | 3.893 (3.156-4.802) | <.0001 | 2.645 (2.080-3.362) | <0.0001 |
|  | 18-24 | 1.512 (1.191-1.920) | 0.0007 | 1.283 (0.993-1.658) | 0.0567 |
| Weekend |  | 0.219 (0.184-0.260) | <.0001 | 0.248 (0.206-0.298) | <0.0001 |
| Laboratory study |  | 329.848 (82.405->999.999) | <.0001 | 65.286 (16.112-264.541) | <0.0001 |
| Imaging study | X-ray | 19.179 (13.002-28.292) | <.0001 | 2.408 (1.595-3.633) | <0.0001 |
|  | CT | 3.671 (3.282-4.106) | <.0001 | 1.971 (1.738-2.234) | <0.0001 |
|  | MRI | 2.061 (1.722-2.467) | <.0001 | 2.557 (2.024-3.229) | <0.0001 |
| Specialty consultation |  | 5.344 (4.602-6.207) | <.0001 | 2.114 (1.784-2.505) | <0.0001 |
| Period | Pre | 1 |  | 1 |  |
|  | Post | 0.514 (0.461-0.574) | <.0001 | 0.433 (0.384-0.489) | <0.0001 |

CI: confidence interval; EMS: emergency medical services; KTAS: Korean Triage and Acuity Scale; ED: emergency department; CT: computed tomography; MRI: magnetic resonance imaging.

Appendix 2. Logistic regression analysis for emergency department length of stay over 24 hours in admitted patients.

| Variables | | Univariable | | Multivariable | |
| --- | --- | --- | --- | --- | --- |
|  |  | Odds ratio (95% CI) | p-value | Odds ratio (95% CI) | p-value |
| Age | -39 | 1 |  | 1 |  |
|  | 40-64 | 1.592 (1.285-1.973) | <0.0001 | 1.303 (1.003-1.648) | 0.0273 |
|  | 65- | 1.602 (1.299-1.976) | <0.0001 | 1.197 (0.946-1.516) | 0.1344 |
| Female |  | 0.824 (0.726-0.934) | 0.0024 | 0.856 (0.747-0.982) | 0.0263 |
| Transfer in |  | 0.932 (0.805-1.080) | 0.3493 |  |  |
| EMS |  | 1.126 (0.993-1.278) | 0.0647 | 1.197 (1.023-1.401) | 0.0249 |
| KTAS | 1 | 1.230 (0.869-1.740) | 0.2432 | 1.406 (0.929-2.128) | 0.1068 |
|  | 2 | 0.750 (0.624-0.902) | 0.0022 | 0.852 (0.682-1.063) | 0.1555 |
|  | 3 | 1 |  | 1 |  |
|  | 4 | 0.804 (0.698-0.926) | 0.0026 | 0.884 (0.730-1.071) | 0.2071 |
|  | 5 | 0.825 (0.606-1.124) | 0.2237 | 0.690 (0.481-0.991) | 0.0444 |
| Complaint category | Gastrointestinal | 1 |  | 1 |  |
|  | General | 0.798 (0.639-0.996) | 0.0008 | 1.380 (1.124-1.694) | 0.0021 |
|  | Neurological | 0.506 (0.265-0.963) | 0.0013 | 0.430 (0.326-0.567) | <0.0001 |
|  | Cardiovascular | 1.371 (1.141-1.647) | 0.0458 | 0.769 (0.598-0.989) | 0.0404 |
|  | Musculoskeletal | 0.581 (0.420-0.802) | 0.001 | 0.438 (0.307-0.625) | <0.0001 |
|  | Respiratory | 0.689 (0.549-0.865) | <0.0001 | 1.472 (1.191-1.818) | 0.0003 |
|  | ENT | 0.607 (0.439-0.840) | 0.0381 | 0.536 (0.268-1.069) | 0.0768 |
|  | Skin | 1.787 (1.487-2.147) | 0.876 | 1.007 (0.535-1.893) | 0.9835 |
|  | Others | 1.047 (0.590-1.856) | 0.0026 | 0.557 (0.391-0.793) | 0.0012 |
| Severe disease |  | 0.784 (0.693-0.887) | 0.0001 | 0.764 (0.661-0.883) | 0.0003 |
| Emergency physician |  | 1.853 (1.620-2.120) | <0.0001 | 1.214 (1.015-1.451) | 0.0335 |
| Area | Monitoring area | 0.801 (0.673-0.954) | 0.0129 | 0.854 (0.666-1.095) | 0.2143 |
|  | Bed area | 0.718 (0.618-0.834) | <0.0001 | 0.840 (0.698-1.010) | 0.0643 |
|  | Chair area | 0.374 (0.302-0.463) | <0.0001 | 0.535 (0.424-0.675) | <0.0001 |
|  | Fast track | 1 |  | 1 |  |
| Time of ED arrival | 0-6 | 1 |  | 1 |  |
|  | 6-12 | 2.264 (1.773-2.890) | <0.0001 | 2.020 (1.534-2.659) | <0.0001 |
|  | 12-18 | 2.542 (1.999-3.233) | <0.0001 | 2.184 (1.662-2.869) | <0.0001 |
|  | 18-24 | 1.321 (1.005-1.737) | 0.0461 | 1.144 (0.854-1.532) | 0.368 |
| Weekend |  | 0.216 (0.177-0.264) | <0.0001 | 0.229 (0.186-0.283) | <0.0001 |
| Laboratory study |  | 7.494 (1.821-30.845) | 0.0053 | 5.098 (1.196-21.732) | 0.0277 |
| Imaging study | X-ray | 2.929 (1.860-4.614) | <0.0001 | 2.306 (1.424-3.734) | 0.0007 |
|  | CT | 1.436 (1.260-1.637) | <0.0001 | 1.572 (1.360-1.818) | <0.0001 |
|  | MRI | 1.215 (0.978-1.509) | 0.079 | 1.790 (1.371-2.338) | <0.0001 |
| Period | Pre | 1 |  |  |  |
|  | Post | 0.464 (0.408-0.528) | <0.0001 | 0.428 (0.372-0.492) | <0.0001 |

CI: confidence interval; EMS: emergency medical services; KTAS: Korean Triage and Acuity Scale; ED: emergency department; CT: computed tomography; MRI: magnetic resonance imaging.

Appendix 3. Generalized linear regression analysis for emergency department length of stay in total patients.

| Variables | | Univariable | | Multivariable | |
| --- | --- | --- | --- | --- | --- |
|  |  | Beta coefficient (95% CI) | p-value | Beta coefficient (95% CI) | p-value |
| Age | -39 | 1 |  | 1 |  |
|  | 40-64 | 194.9 (176.2-213.7) | <0.0001 | 42.7 (25.5-59.9) | <0.0001 |
|  | 65- | 323.8 (304.5-343.1) | <0.0001 | 38.1 (19.1-57.1) | <0.0001 |
| Female |  | -101.1 (-116.8--85.3) | <0.0001 | -32.4 (-46.3--18.5) | <0.0001 |
| Transfer in |  | 257.0 (232.8-281.2) | <0.0001 | 18.4 (-3.8-40.6) | 0.1039 |
| EMS |  | 243.0 (224.8-261.2) | <0.0001 | 25.3 (6.7-44.0) | 0.0079 |
| KTAS | 1 | 178.7 (115.7-241.6) | <0.0001 | 135.5 (72.5-198.6) | <0.0001 |
|  | 2 | 100.3 (69.8-130.9) | <0.0001 | 43.7 (14.0-73.4) | 0.0039 |
|  | 3 | 1 |  | 1 |  |
|  | 4 | -253.1 (-271.6--234.6) | <0.0001 | -72.1 (-92.6--51.7) | <0.0001 |
|  | 5 | -374.2 (-400.6--347.7) | <0.0001 | -82.4 (-111.1--53.6) | <0.0001 |
| Complaint category | Gastrointestinal | 1 |  | 1 |  |
|  | General | -25.7 (-51.0--0.4) | 0.0468 | 61.7 (38.4-85.0) | <0.0001 |
|  | Neurological | -104.2 (-130.8--77.7) | <0.0001 | -251.7 (-278.6--224.9) | <0.0001 |
|  | Cardiovascular | -48.0 (-77.2--18.8) | 0.0013 | -116.2 (-143.9--88.6) | <0.0001 |
|  | Musculoskeletal | -202.1 (-233.3--170.9) | <0.0001 | -66.6 (-97.6--35.7) | <0.0001 |
|  | Respiratory | 365.9 (333.7-398.1) | <0.0001 | 215.9 (186.0-245.8) | <0.0001 |
|  | ENT | -291.4 (-325.9--256.9) | <0.0001 | -91.2 (-124.9--57.4) | <0.0001 |
|  | Skin | -302.1 (-336.8--267.3) | <0.0001 | -30.6 (-65.2-4.0) | 0.0827 |
|  | Others | -165.3 (-195.9--134.8) | <0.0001 | -93.0 (-121.9--64.1) | <0.0001 |
| Severe disease |  | 478.7 (459.2-498.1) | <0.0001 | 162.1 (141.6-182.5) | <0.0001 |
| Emergency physician |  | 256.6 (240.9-272.3) | <0.0001 | 24.9 (6.8-43.1) | 0.0072 |
| Area | Monitoring area | 534.9 (505.5-564.3) | <0.0001 | 105.7 (70.4-141.0) | <0.0001 |
|  | Bed area | 367.9 (347.4-388.4) | <0.0001 | 84.7 (62.0-107.3) | <0.0001 |
|  | Chair area | 77.8 (57.7-97.9) | <0.0001 | -21.6 (-41.8--1.5) | 0.0353 |
|  | Fast track | 1 |  | 1 |  |
| Time of ED arrival | 0-6 | 1 |  | 1 |  |
|  | 6-12 | 158.3 (134.7-181.9) | <0.0001 | 42.1 (19.3-64.9) | 0.0003 |
|  | 12-18 | 179.9 (156.6-203.3) | <0.0001 | 64.9 (42.1-87.6) | <0.0001 |
|  | 18-24 | 72.7 (48.1-97.3) | <0.0001 | 44.2 (22.4-65.9) | <0.0001 |
| Weekend |  | -168.9 (-185.7--152.2) | <0.0001 | -110.2 (-125.4—95.0) | <0.0001 |
| Laboratory study |  | 416.2 (399.6-432.8) | <0.0001 | 141.1 (118.8-163.4) | <0.0001 |
| Imaging study | X-ray | 362.0 (344.1-379.9) | <0.0001 | 52.3 (29.9-74.6) | <0.0001 |
|  | CT | 354.3 (338.7-369.9) | <0.0001 | 195.6 (179.7-211.5) | <0.0001 |
|  | MRI | 239.8 (204.9-274.7) | <0.0001 | 219.9 (185.6-254.2) | <0.0001 |
| Specialty consultation |  | 389.7 (374.6-404.8) | <0.0001 | 178.0 (162.0-194.0) | <0.0001 |
| Period | Pre | 1 |  | 1 |  |
|  | Post | -86.1 (-101.9--70.4) | <0.0001 | -99.3 (-113.1--85.4) | <0.0001 |

CI: confidence interval; EMS: emergency medical services; KTAS: Korean Triage and Acuity Scale; ED: emergency department; CT: computed tomography; MRI: magnetic resonance imaging.

Appendix 4. Generalized linear regression analysis for emergency department length of stay in admitted patients.

| Variables | | Univariable | | Multivariable | |
| --- | --- | --- | --- | --- | --- |
|  |  | Beta coefficient (95% CI) | p-value | Beta coefficient (95% CI) | p-value |
| Age | -39 | 1 |  | 1 |  |
|  | 40-64 | 178.6 (95.8-261.3) | <0.0001 | 64.5 (-15.1-144.1) | 0.1123 |
|  | 65- | 188.4 (108.0-268.9) | <0.0001 | 43.2 (-36.9-123.4) | 0.2908 |
| Female |  | -66.6 (-119.3--13.9) | 0.0132 | -39.2 (-88.9-10.6) | 0.1231 |
| Transfer in |  | -49.8 (-111.5-11.8) | 0.1132 | -87.8 (-147.6—28.0) | 0.0040 |
| EMS |  | 77.8 (24.0-131.5) | 0.0046 | 59.3 (0.3-118.3) | 0.0489 |
| KTAS | 1 | 263.4 (105.2-421.5) | 0.0011 | 282.6 (121.2-444.0) | 0.0006 |
|  | 2 | -63.2 (-139.2-12.9) | 0.1034 | 4.3 (-76.5-85.0) | 0.9174 |
|  | 3 | 1 |  | 1 |  |
|  | 4 | -107.8 (-167.8--47.8) | 0.0004 | -82.3 (-151.8--12.8) | 0.0203 |
|  | 5 | -132.7 (-261.6--3.8) | 0.0436 | -161.4 (-292.5--30.3) | 0.0159 |
| Complaint category | Gastrointestinal | 1 |  | 1 |  |
|  | General | 159.4 (80.1-238.6) | <0.0001 | 156.1 (77.6-234.6) | <0.0001 |
|  | Neurological | -209.6 (-296.1--123.0) | <0.0001 | -420.9 (-515.2--326.5) | <0.0001 |
|  | Cardiovascular | -127.2 (-214.5--39.8) | 0.0043 | -137.3 (-227.0--47.7) | 0.0027 |
|  | Musculoskeletal | -215.0 (-330.2--99.8) | 0.0003 | -262.8 (-379.1--146.5) | <0.0001 |
|  | Respiratory | 366.7 (284.9-448.4) | <0.0001 | 281.9 (198.8-365.1) | <0.0001 |
|  | ENT | -246.2 (-461.1--31.4) | 0.0247 | -182.9 (-389.6-23.9) | 0.0830 |
|  | Skin | -29.1 (-268.0-209.9) | 0.8115 | 24.9 (-205.9-255.8) | 0.8323 |
|  | Others | -196.3 (-313.4--79.2) | 0.0010 | -184.2 (-299.7--68.7) | 0.0018 |
| Severe disease |  | -52.5 (-104.8--0.2) | 0.0493 | -66.0 (-119.7--12.4) | 0.0159 |
| Emergency physician |  | 149.7 (96.2-203.2) | <0.0001 | 25.4 (-38.7-89.5) | 0.4379 |
| Area | Monitoring area | 26.0 (-50.5-102.5) | 0.5051 | -47.4 (-141.7-47.0) | 0.3255 |
|  | Bed area | -61.9 (-127.2-3.3) | 0.0629 | -54.7 (-124.3-14.9) | 0.1234 |
|  | Chair area | -250.4 (-329.5--171.3) | <0.0001 | -141.4 (-218.8--64.0) | 0.0003 |
|  | Fast track | 1 |  | 1 |  |
| Time of ED arrival | 0-6 | 1 |  | 1 |  |
|  | 6-12 | 117.3 (29.5-205.0) | 0.0089 | 73.4 (-16.1-162.8) | 0.1079 |
|  | 12-18 | 133.0 (46.8-219.2) | 0.0025 | 84.1 (-5.2-173.3) | 0.0649 |
|  | 18-24 | 161.7 (65.6-257.9) | 0.0010 | 128.8 (37.0-220.5) | 0.0060 |
| Weekend |  | -412.7 (-471.2--354.2) | <0.0001 | -375.6 (-433.9--317.4) | <0.0001 |
| Laboratory study |  | 577.9 (301.9-853.9) | <0.0001 | 268.8 (1.3-536.2) | 0.0489 |
| Imaging study | X-ray | 481.6 (346.5-616.7) | <0.0001 | 326.4 (193.3-459.4) | <0.0001 |
|  | CT | 227.8 (174.3-281.2) | <0.0001 | 228.1 (175.7-280.5) | <0.0001 |
|  | MRI | 134.9 (39.2-230.5) | 0.0057 | 329.8 (230.2-429.4) | <0.0001 |
| Period | Pre | 1 |  | 1 |  |
|  | Post | -306.1 (-357.8--254.4) | <0.0001 | -310.9 (-360.6--261.3) | <0.0001 |

CI: confidence interval; EMS: emergency medical services; KTAS: Korean Triage and Acuity Scale; ED: emergency department; CT: computed tomography; MRI: magnetic resonance imaging.
